# Temporal Dynamics of Daily Sleep, Mood, and Cognition in NHS Shift Workers: A Digital Experience Sampling (ESM) Study

**DOI:** 10.64898/2026.08.27.26361516

**Authors:** Robert Hickman, Dan W. Joyce, Nicholas Gray, Adam Hampshire, Peter J. Hellyer, Ziyuan Cai, Sukhi Shergill, Teresa C. D’Oliveira

**Author notes:** Please note that the indicated authors are co-senior authors on this paper (Sukhi Shergill, Teresa C. D’Oliveira).

## Abstract

**Background:** Sleep, mood, and affective states are mutually connected. There is a paucity of studies, however, that have considered bidirectional relationships between daily sleep-affective dyads in naturalistic settings, particularly for shift workers.

**Objective:** To evaluate the dynamic and temporal interplay of daily smartphone-based self-reported sleep measurements, dimensions of affective experience and cognitive processing in UK shift working nurses.

**Methods:** The *EClocker Study* prospectively monitored 102 National Health Service (NHS) nurses (aged 25-61 years, 83.3% female) working standard (day shift) and non-standard (fast rotating shifts) schedules over a two-week period. Smartphone-based Experience Sampling Methodology (ESM) recorded daily sleep, mood, momentary affect and cognitive attentional functioning. Self-reported burnout, emotional dysregulation, emotion reactivity and affective dimensions (positive and negative) were also collected.

**Findings:** Overall, NHS nurses reported a high prevalence of depressive symptoms, stress, burnout and sleep-circadian rhythm disturbances. Generalised Additive Modelling (GAMs) revealed that NHS nurses’ higher perceived sleep quality predicted better next-day mood state, while better daytime mood was associated with reduced sleep onset latency, such that participants reported falling asleep faster. In contrast, daytime mood or affect (positive and negative) had no substantial, direct impact on nurses’ subjective sleep parameters (sleep quality, sleep duration, sleep efficiency). Exposure to fast rotating night shifts across the two-week study was associated with more frequent response errors on a Choice Reaction Time (CRT) cognitive task, while daytime somnolence did not adversely influence nurses’ momentary reaction time speeds or attentional function.

**Conclusions:** Clinically relevant sleep impairments, insomnia-related symptoms, elevated stress, and poor mood were pervasive in a sample of UK NHS nurses, regardless of shift type. Sleep quality impacted next-day mood and daytime mood impacted sleep latency, while rotating shifts led to an increase in cognitive errors. Recognising the impact of shiftwork and designing interventions to promote better sleep quality offer potential to enhance mood and performance in healthcare professionals.

**Clinical implications:** We need to implement and evaluate interventions that regularise sleep patterns and promote sleep quality to alleviate mood symptoms among frontline NHS shift workers.

## WHAT IS ALREADY KNOWN ON THIS TOPIC

- Previous ambulatory monitoring studies have shown strong temporal and reciprocal sleep-affect connections. There remains an underexplored research gap, however, in monitoring daily sleep-affect dyads in shift workers (especially healthcare staff) who are vulnerable to circadian misalignment and sleep instabilities.

## WHAT THIS STUDY ADDS

- The *EClocker Study* used smartphone-based Experience Sampling Methods (ESM) to monitor and tease apart the bidirectional relationships between daily sleep patterns and affective experiences in a cohort of NHS healthcare staff.

## HOW THIS STUDY MIGHT AFFECT RESEARCH, PRACTICE OR POLICY

- Maintaining sufficient sleep quality and sleep satisfaction emerged as a core sleep index which may promote better mood symptoms. Prioritising sleep quality could thus represent a modifiable, cost-effective therapeutic target for improving mood stability in NHS shift workers.

## BACKGROUND

Sleep health is considered an essential indicator and proxy of overall health (1) but is often compromised in shift workers. The National Health Service (NHS) is the largest employer in Europe and has the highest number of UK shift workers. Nurses and midwives form the second highest proportion of UK night shift staff and are prone to poor sleep, excessive daytime sleepiness, and reduced sleep opportunities (2, 3). Shift work and long hours are common in the NHS and recognised by the Royal College of Nursing (RCN), American Academy of Nursing (AAN) and the British Medical Association (BMA) as posing a profound risk for widespread fatigue and chronic sleep-circadian disruption.

A number of reviews have identified poor acute and chronic sleep in nurses (4) and the demonstrable negative health impacts of sleep loss connected to shift work (5). Insomnia and poor sleep quality, for example, are endemic among nursing and healthcare staff and are more prevalent relative to the general population or other professions (3, 4). Circadian misalignment through night shift work confers vulnerability to poor mental health, increased sickness absence, negatively impacts mood, and contributes to increased risk onset of psychiatric disorders (6–11). Sleep disturbances amplify negative affect, diminish positive affect, compromise emotional regulatory processes, emotional empathy, emotional reactivity and is linked to detrimental next-day mood (12, 13).

Few Experience Sampling Method (ESM) studies have collected comprehensive data with shift workers (14), with only six studies to our knowledge utilising prospective ambulatory designs to examine day-to-day sleep-mood associations among healthcare shift workers (12). Four of these identified studies were with medical residents and only two with nursing staff. None of these studies with healthcare staff were situated in the UK.

## OBJECTIVE

The *EClocker Study* addressed this research gap by utilising smartphone-based ESM to examine the relationship between sleep characteristics, daily mood, momentary affective states and cognitive attentional performance amongst UK NHS healthcare staff on two different shift patterns.

## METHODS

### Participants and study design

Sample characteristics are presented in **Table 1** and were demographically comparable to the wider UK nursing and midwifery workforce. NHS staff were fully qualified and registered NHS nurses with the Nursing and Midwifery Council (NMC), working at three London NHS Foundation Trusts. One hundred and two NHS nurses were enrolled between March 2022 and June 2023, of which 49 were day shift workers (DSW; 48%) and 53 were fast rotating shift workers (NSW; 52%) (**Supplemental File 1, Figure S1**). NHS nurses working fast rotating shifts from *EClocker Phase 1* could opt-in to participate in a pilot (non-randomised) sleep aid intervention in *EClocker Phase 2*.

**Table 1.**
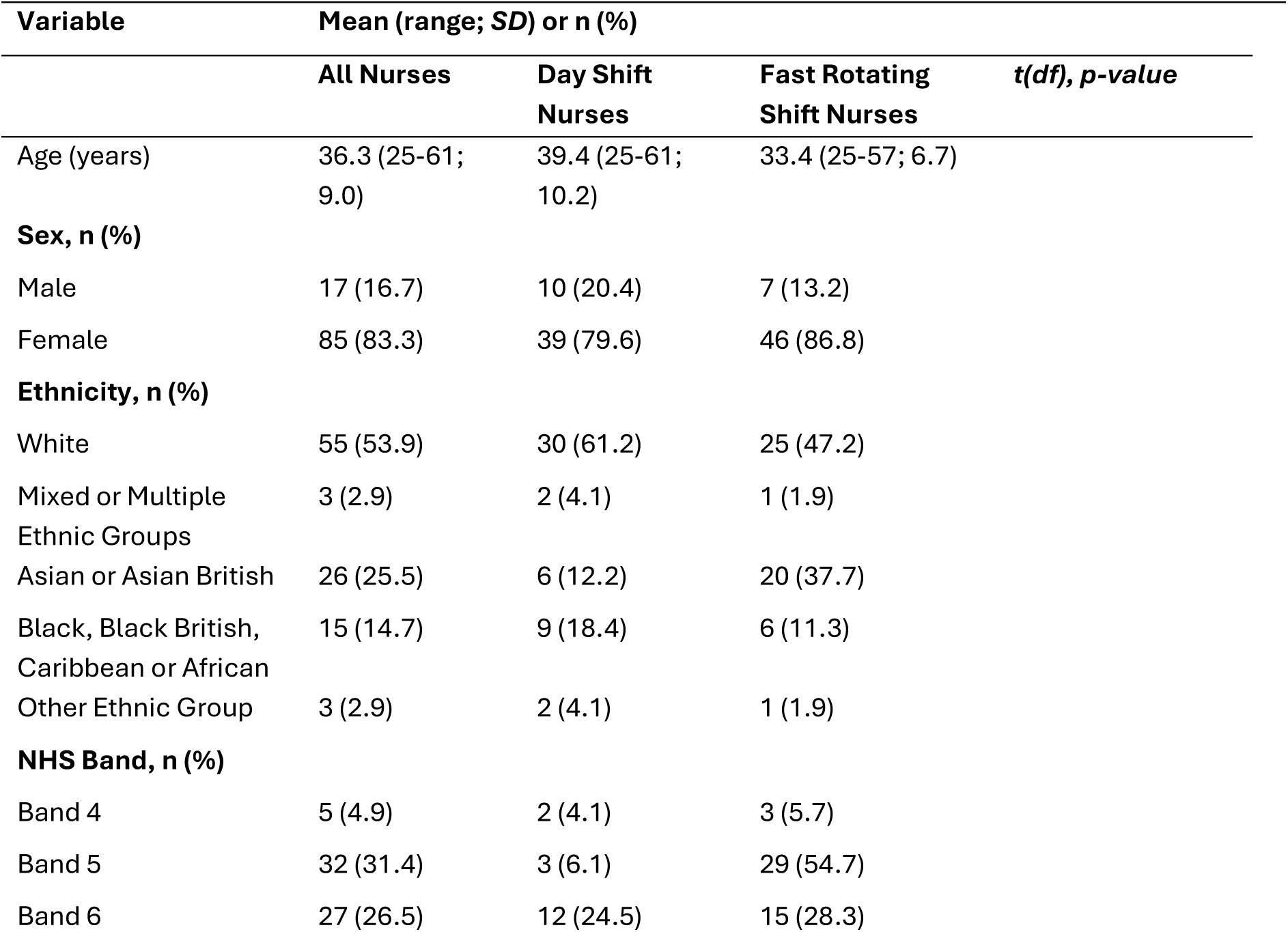
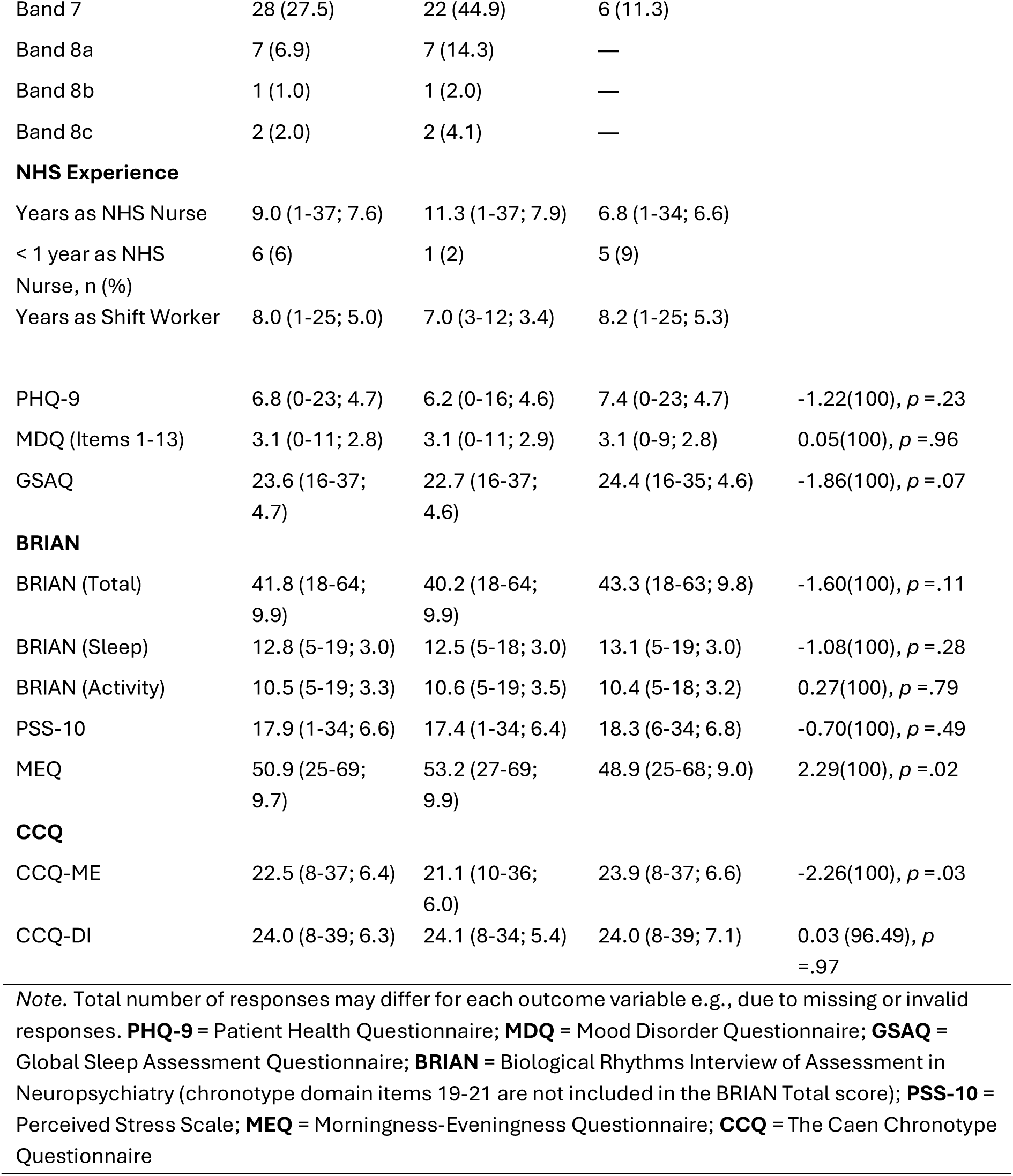
Characteristics of NHS nurses at study entry (baseline screening responses) who participated in the *EClocker Phase 1* two-week data collection. All Nurses (N = 102), Day Shift Nurses (N = 49), Fast Rotating Shift Nurses (N = 53)

| Variable | Mean (range; SD) or n (%) |  |  | t(df), p-value |
| --- | --- | --- | --- | --- |
|  | All Nurses | Day Shift Nurses | Fast Rotating Shift Nurses |  |
| Age (years) | 36.3 (25-61; 9.0) | 39.4 (25-61; 10.2) | 33.4 (25-57; 6.7) |  |
| **Sex, n (%)** |  |  |  |  |
| Male | 17 (16.7) | 10 (20.4) | 7 (13.2) |  |
| Female | 85 (83.3) | 39 (79.6) | 46 (86.8) |  |
| **Ethnicity, n (%)** |  |  |  |  |
| White | 55 (53.9) | 30 (61.2) | 25 (47.2) |  |
| Mixed or Multiple Ethnic Groups | 3 (2.9) | 2 (4.1) | 1 (1.9) |  |
| Asian or Asian British | 26 (25.5) | 6 (12.2) | 20 (37.7) |  |
| Black, Black British, Caribbean or African | 15 (14.7) | 9 (18.4) | 6 (11.3) |  |
| Other Ethnic Group | 3 (2.9) | 2 (4.1) | 1 (1.9) |  |
| **NHS Band, n (%)** |  |  |  |  |
| Band 4 | 5 (4.9) | 2 (4.1) | 3 (5.7) |  |
| Band 5 | 32 (31.4) | 3 (6.1) | 29 (54.7) |  |
| Band 6 | 27 (26.5) | 12 (24.5) | 15 (28.3) |  |
| Band 7 | 28 (27.5) | 22 (44.9) | 6 (11.3) |  |
| Band 8a | 7 (6.9) | 7 (14.3) | — |  |
| Band 8b | 1 (1.0) | 1 (2.0) | — |  |
| Band 8c | 2 (2.0) | 2 (4.1) | — |  |
| **NHS Experience** |  |  |  |  |
| Years as NHS Nurse | 9.0 (1-37; 7.6) | 11.3 (1-37; 7.9) | 6.8 (1-34; 6.6) |  |
| < 1 year as NHS Nurse, n (%) | 6 (6) | 1 (2) | 5 (9) |  |
| Years as Shift Worker | 8.0 (1-25; 5.0) | 7.0 (3-12; 3.4) | 8.2 (1-25; 5.3) |  |
| PHQ-9 | 6.8 (0-23; 4.7) | 6.2 (0-16; 4.6) | 7.4 (0-23; 4.7) | -1.22(100), $p = .23$ |
| MDQ (Items 1-13) | 3.1 (0-11; 2.8) | 3.1 (0-11; 2.9) | 3.1 (0-9; 2.8) | 0.05(100), $p = .96$ |
| GSAQ | 23.6 (16-37; 4.7) | 22.7 (16-37; 4.6) | 24.4 (16-35; 4.6) | -1.86(100), $p = .07$ |
| **BRIAN** |  |  |  |  |
| BRIAN (Total) | 41.8 (18-64; 9.9) | 40.2 (18-64; 9.9) | 43.3 (18-63; 9.8) | -1.60(100), $p = .11$ |
| BRIAN (Sleep) | 12.8 (5-19; 3.0) | 12.5 (5-18; 3.0) | 13.1 (5-19; 3.0) | -1.08(100), $p = .28$ |
| BRIAN (Activity) | 10.5 (5-19; 3.3) | 10.6 (5-19; 3.5) | 10.4 (5-18; 3.2) | 0.27(100), $p = .79$ |
| PSS-10 | 17.9 (1-34; 6.6) | 17.4 (1-34; 6.4) | 18.3 (6-34; 6.8) | -0.70(100), $p = .49$ |
| MEQ | 50.9 (25-69; 9.7) | 53.2 (27-69; 9.9) | 48.9 (25-68; 9.0) | 2.29(100), $p = .02$ |
| **CCQ** |  |  |  |  |
| CCQ-ME | 22.5 (8-37; 6.4) | 21.1 (10-36; 6.0) | 23.9 (8-37; 6.6) | -2.26(100), $p = .03$ |
| CCQ-DI | 24.0 (8-39; 6.3) | 24.1 (8-34; 5.4) | 24.0 (8-39; 7.1) | 0.03 (96.49), $p = .97$ |

*Note.* Total number of responses may differ for each outcome variable e.g., due to missing or invalid responses. **PHQ-9** = Patient Health Questionnaire; **MDQ** = Mood Disorder Questionnaire; **GSAQ** = Global Sleep Assessment Questionnaire; **BRIAN** = Biological Rhythms Interview of Assessment in Neuropsychiatry (chronotype domain items 19-21 are not included in the BRIAN Total score); **PSS-10** = Perceived Stress Scale; **MEQ** = Morningness-Eveningness Questionnaire; **CCQ** = The Caen Chronotype Questionnaire

Acceptability and perceived effectiveness of the sleep intervention was assessed in this second phase (*EClocker Phase 2* data not reported here). Additional details of the *EClocker Study* protocol are available in **Supplemental File 1**, with all measures referenced and described in full.

### Screening

NHS nurses completed an online screening questionnaire (Qualtrics) prior to study enrolment. Depressive symptoms were assessed using the Patient Health Questionnaire (PHQ-9), bipolar disorder with the Mood Disorder Questionnaire (MDQ) and stress levels with the Perceived Stress Scale (PSS-10). Sleep disorders were measured using the Global Sleep Assessment Questionnaire (GSAQ) and biological rhythm disruption across the domains of sleep, activity, social and eating patterns with the Biological Rhythms Interview of Assessment in Neuropsychiatry (BRIAN). Chronotype was assessed with the Morningness-Eveningness Questionnaire (MEQ) and The Caen Chronotype Questionnaire (CCQ) for a circadian amplitude dimension of diurnal variation. Demographic, lifestyle, health data, anthropometric measurements, NHS employment history and work schedule characteristics were also collected during screening.

Inclusion criteria were (1) NMC registered NHS nurses working standard (day shift) or non-standard (fast rotating shifts) at participating London NHS Trusts and (2) aged ≥21 years. Exclusion criteria were (1) any mood disorder or sleep disorder diagnosis; alcohol / other substance dependence (excluding caffeine / nicotine) in the last 12 months, (2) illicit substance / psychostimulant use in the last 6 months, (3) pregnancy, (4) receiving a sleep intervention / treatment (including being a participant in a clinical research study) or a medication indicated for, or taken to improve, sleep (e.g., hypnotics, sedatives, melatonin) in the last 12 months.

### Daily Smartphone-based ESM Monitoring

Nurses used the ExpiWell smartphone app (a commercial experience sampling platform; https://www.expiwell.com/) to complete daily sleep and affective diaries and respond to momentary ESM measures over two-weeks (**Supplemental File 1, Table S1** for full task items). The *EClocker Study* sampling period was in line with prior research (12) and consistent with the International Classification of Sleep Disorders (ICSD-3) and American Academy of Sleep Medicine (AASM) guidelines to capture sleep and daily function variability. Push-notifications and survey scheduling were designed to be minimally burdensome and tailored for each nurse according to their shift schedule (**Supplemental File 1, Table S2**). As part of the larger *EClocker Study*, ankle-worn actigraphs (ActiGraph wGT3X-BT devices; ActiGraph LLC, Pensacola, USA) were also worn concurrently (device data not reported here).

### Sleep Diary

An adapted version of the Consensus Sleep Diary (CSD) was administered daily via ExpiWell. Primary CSD sleep parameters included: time into-out of bed, total sleep time (TST), sleep onset latency (SOL), sleep efficiency (SE), number and duration of awakenings and sleep quality (SQ) which was rated from 1 (very poor) to 5 (very good). All CSD sleep-wake times were recorded to the nearest 15-minute interval. An open-ended section enabled free response entries. Definitions and calculations of CSD sleep indices are outlined in **Supplemental File 1, Table S3.** At the end of the sleep diary, mood valence was rated on a scale from 1 (Lowest) to 10 (Highest). Nurses were prompted to complete the sleep diary preferably within one hour of getting out of bed.

Sleep diaries were scheduled early in the morning (05:30 AM) and were available for the remainder of that day to maximise completion rates.

### Affective Events Diary

Nurses recorded daily events and experiences which had an emotional impact (positive or negative) on them via an adapted Affective Events Diary (AED) on ExpiWell. The mood valence item (as per the sleep diary) was rated from 1 (Lowest) to 10 (Highest). Self-report items related to daily napping, alcohol, caffeine intake, physical activity and sleep medication were also reported.

### Affect and Sleepiness

Momentary affect and sleepiness were collected at the same ESM push-notification. Ten adjectives (four positive and six negative) were rated using 7-point Likert items (anchored at 0 = not at all, to 7 = very much) (e.g., ‘Right now, I feel down’). Sleepiness was captured with the Karolinska Sleepiness Scale (KSS).

### Attentional Function

The Choice Reaction Time (CRT) assessed daily attentional function, reaction time performance, and processing speed (**Supplemental File 1, Figure S2**). The CRT is a short 2-minute task developed by Cognitron (H2 Cognitive Design Ltd) and was delivered via a secure link on ExpiWell.

## Retrospective Measures

At the end of the two-week monitoring phase, a battery of online measures (Qualtrics) was administered. Sleep disturbance and quality was assessed with the Pittsburgh Sleep Quality Index (PSQI). Emotion dysregulation was evaluated with the Difficulties in Emotion Regulation Scale Short Form (DERS-SF), emotion regulation strategies with the Emotional Regulation Strategy Questionnaire at Work (ERSQ-W), and trait emotional reactivity with the Emotion Reactivity Scale (ERS). Affect was assessed with the Positive and Negative Affect Schedule (PANAS-GEN) and burnout symptoms with the Maslach Burnout Inventory – Human Services Survey for Medical Personnel (MBI-HSS MP). NHS nurses perceived work schedule control and recovery activities were also captured.

## Statistical Analyses

Quantitative analyses were undertaken in Python for all data cleaning, pre-processing, and General Additive Models (GAMs) (15). All procedures, including an *a priori* sample size calculation, are reported in **Supplemental File 1**. Descriptive summaries of baseline screening characteristics (**Table 1**) including parametric t-tests were processed in IBM SPSS Statistics (V.29).

GAM models assessed bidirectional relationships between day-to-day ESM variables for sleep, mood, affect (positive and negative) and cognitive attentional function. GAM models were produced for the main dependent variables being sleep parameters (TST, SQ, SOL, SE) identified from reviewed literature (12) which are core sleep dimensions critical for optimal health and functioning. Covariates were based on well-established shift work adaptability and individual factors. Each GAM was adjusted to control for the potential confounding effect of age, baseline depression severity (PHQ-9), chronotype (MEQ), emotion dysregulation (DERS-SF) and fast rotating shift work status (NSW).

Empirical Distribution Functions (ECDF) and Kolmogorov–Smirnov Tests (Two-Sided) for total sleep time (TST) and sleep efficiency (SE) are reported in **Supplemental File 1**.

These tests were performed to ensure normality assumptions were met and to observe group distributions and residuals. As GAMs make use of smoothing splines, the optimal spline points were treated as hyper-parameters to be estimated.

## FINDINGS

### Clinical Characteristics

Score interpretation threshold values for *EClocker Study* measures are provided in **Supplementary File 1.** Day shift nurses and nurses working fast rotating shifts showed no significant differences in baseline depressive symptoms, sleep disorder symptoms, circadian rhythm metrics or stress levels (**Table 1**). NHS nurses on fast rotating shifts, however, had lower MEQ and higher CCQ-ME scores on average than day shift nurses, indicating a stronger evening-type chronotype preference (**Table 1**).

### Sleep Disorder Symptoms

Nearly all NHS nurses (98.0%) experienced one potential sleep disorder symptom to some degree (*at least sometimes*) over the month prior to study entry. Symptoms were based on GSAQ Items 1-9 (excluding work-related Item 4) and responses were categorised as a binary variable representing *‘at least sometimes’* if participants responded to any item as ‘*Sometimes’, ‘Usually’,* or *‘Always*’ in line with prior studies. Insomnia symptoms (difficulty initiating and maintaining sleep) were experienced *at least sometimes* by 91.2% of NHS nurses in the past month. Insomnia-related symptoms (excessive daytime sleepiness) and bothersome daytime sleepiness or sleep difficulties were reported *at least sometimes* by 60% of nurses, while 77.5% of

NHS nurses reported insufficient sleep (*at least sometimes*) due to work or other activities.

### Sleep Quality and Efficiency

Poor subjective sleep quality and potentially clinically relevant sleep impairment was indicated for 77.6% of NHS nurses (PSQI Global scores ≥5). Across all nurses, 43% reported sleep efficiency (SE) of less than 85% over the past month (PSQI SE <85% from Items 1,3, 4). Poor sleep efficiency (SE <85%) was also observed with day-to-day CSD sleep diary data over two-weeks; on average SE across NHS nurses was 82% and lower for nurses on fast rotating shifts at 80%. These SE scores (<85%) may be indicative of problematic sleep disturbance or insomnia symptoms and are below optimal sleep efficiency ranges of 85-90%.

### Circadian Rhythm Disturbance

Over half of NHS nurses (52.9%) had BRIAN Total scores (≥40) indicating potentially severe circadian rhythm dysregulation. BRIAN Total scores (≥40) have previously identified Delayed Sleep-Wake Phase Disorder (DSWPD).

### Depressive Symptoms

Over a third of NHS nurses (36.3%) reported PHQ-9 total scores (8-11 or higher) that met cut-off symptom thresholds for possible major depression. A notable proportion of nurses (29.4%) had ‘yellow flag’ PHQ-9 depressive symptom scores of 10 or greater which are considered potentially clinically significant. At study entry, nearly 70% of NHS nurses also reported (*at least sometimes*) feeling sad or anxious over the past month (GSAQ Item 15) and had worries which disturbed their sleep (GSAQ Item 12).

### Stress and Burnout

Around three quarters (74.5%) of NHS nurses reported moderate or high stress levels (PSS-10) and nearly 1 in 4 NHS nurses (24.7%) met criteria for burnout (MBI-HSS MP).

### General Additive Models (GAMs)

Sampling from 102 NHS nurses resulted in a total of 1439 days of smartphone-based ESM observations with sleep, mood, and affect outcome scores summarised in **Supplemental File 1, Table S7**. To compare day (DSW) and rotating night shift (NSW) nurses, only CSD sleep diaries completed following a day shift or non-working day (no contracted work hours) were included in GAM models.

### Daily Mood on Sleep

In total, 691 daily smartphone-based ESM observations were included in the mood- sleep GAM models. Higher daily mood (average mood score derived from all daily mood ratings) decreased sleep latency (SOL, negative coefficient -2.76; **Table 2)** and had little impact on sleep duration (TST), sleep efficiency (SE), or sleep quality (SQ) across all NHS nurses.

**Table 2.** GAM. Sleep Latency (SOL) ∼ Mood + NSW + Age + PHQ-9 + MEQ

|  | **Coefficient** | **p-value** | **95% CI** |
| --- | --- | --- | --- |
| Intercept | 75.247 | 0.000 | [45.722, 104.771] |
| Mood | -2.761 | 0.019 | [-5.067, -0.456] |
| NSW | 3.510 | 0.001 | [1.410, 5.611] |
| Age | 0.357 | 0.036 | [0.024, 0.691] |
| PHQ-9 | 0.230 | 0.467 | [-0.390, 0.849] |
| MEQ | -0.332 | 0.124 | [-0.756, 0.091] |

**SOL** = Sleep Latency; **NSW** = Exposure to fast rotating shifts; **PHQ-9** = Patient Health Questionnaire; **MEQ** = Morningness-Eveningness Questionnaire

### Affect on Sleep

In total, 1076 smartphone-based ESM observations were included in the affect-sleep GAM models. Emotion regulation (DERS-SF) was an independent trait that was positively associated with sleep parameters (TST, SQ, SOL, SE) and both positive and negative affect dimensions. There was no impact of positive affect (PA) or negative affect (NA) scores on sleep variables (TST, SQ, SOL, SE).

### Sleep on Next-Day Mood

In total, 785 smartphone-based ESM observations were included in the sleep-mood GAM model. Sleep efficiency (SE) was removed from the model to avoid collinearity with other sleep variables. Sleep quality (SQ) was associated with next-day mood, with no impact related to sleep time or latency. Higher perceived nighttime sleep quality (SQ; **Table 3**) predicted higher (average) mood the following day (coefficient = 1.02, CI [0.70, 1.33], p-value = 0.00).

**Table 3.** GAM. Mood ∼ TST + SQ + SOL + NSW + Age + PHQ-9 + MEQ + DERS-SF

|  | **Coefficient** | **p-value** | **95% CI** |
| --- | --- | --- | --- |
| Intercept | 2.476 | 0.000 | [1.294, 3.658] |
| TST | 0.056 | 0.449 | [-0.089, 0.202] |
| SQ | 1.017 | 0.000 | [0.705, 1.329] |
| SOL | -0.006 | 0.065 | [-0.011, 0.000] |
| NSW | 0.056 | 0.367 | [-0.066, 0.178] |
| Age | 0.052 | 0.000 | [0.030, 0.075] |
| PHQ-9 | -0.100 | 0.000 | [-0.135, -0.066] |
| MEQ | -0.015 | 0.280 | [-0.043, 0.013] |
| DERS-SF | 0.020 | 0.091 | [-0.003, 0.043] |

**TST** = Total Sleep Time; **SQ** = Sleep Quality; **SOL** = Sleep Latency; **NSW** = Exposure to fast rotating shifts; **PHQ-9** = Patient Health Questionnaire; **MEQ** = Morningness-Eveningness Questionnaire; **DERS-SF** = Difficulties in Emotion Regulation Scale Short Form

### Sleep on Next-Day Affect

In total, 735 smartphone-based ESM observations were included in the sleep-affect GAM models. Sleep parameters (TST, SQ, SOL) did not impact next-day positive affect (PA) scores. Higher perceived sleep quality was associated with a small magnitude increase in negative affect (NA) scores (coefficient = 0.19, CI [0.03, 0.36], p-value = 0.02).

### Sleep on Next-Day Cognition

In total, 476 smartphone-based ESM observations were included in the sleep-cognition GAM models. There was no impact of daily sleepiness (KSS) on CRT attention or processing speed outcomes: Number of Incorrect Trials (i.e., wrong responses only, excluding timeouts / non-responses); Median Reaction Time for all trials; and CRT Summary scores (mean reaction time for correct trials).

However, exposure to fast rotating shifts (NSW) during the study appeared to worsen cognitive performance (**Table 4**), with a positive association with the number of incorrect CRT trial responses (coefficient = 1.56, CI [0.69, 2.42], p-value = 0.00).

**Table 4.** GAM. CRT Number Incorrect Trials ∼ Sleepiness + SQ + SOL+ NSW + Age + PHQ-9 +MEQ + DERS-SF

|  | **Coefficient** | **p-value** | **95% CI** |
| --- | --- | --- | --- |
| Intercept | 6.722 | 0.052 | [-0.068, 13.512] |
| Sleepiness | 0.064 | 0.791 | [-0.408, 0.536] |
| SQ | 1.296 | 0.160 | [-0.513, 3.106] |
| SOL | -0.027 | 0.159 | [-0.064, 0.010] |
| NSW | 1.555 | 0.000 | [0.695, 2.416] |
| Age | 0.168 | 0.017 | [0.030, 0.306] |
| PHQ-9 | -0.002 | 0.987 | [-0.264, 0.260] |
| MEQ | -0.005 | 0.958 | [-0.175, 0.166] |
| DERS-SF | -0.100 | 0.171 | [-0.242, 0.043] |

**CRT** = Choice Reaction Time Task; **SQ** = Sleep Quality; **SOL** = Sleep Latency; **NSW** = Exposure to fast rotating shifts; **PHQ-9** = Patient Health Questionnaire; **MEQ** = Morningness-Eveningness Questionnaire; **DERS-SF** = Difficulties in Emotion Regulation Scale Short Form

## DISCUSSION

To our knowledge, this is the first study incorporating digital ESM sampling to monitor reciprocal associations of nightly sleep and daily affective experiences (mood or affect) in a cohort of NHS healthcare shift workers. Better daytime mood or affect (positive and negative) did not impact subjective nightly sleep quality (SQ), sleep duration (TST), or sleep efficiency (SE). Better (average) daytime mood, however, was associated with shorter sleep onset latency (SOL). In other words, better overall mood during the day, resulted in NHS nurses falling asleep faster. Higher perceived nighttime sleep quality (SQ) also predicted better mood the following day. Emotion regulation ability (DERS-SF) emerged as an innate, trait-based measure that was positively associated with sleep (TST, SQ, SOL, SE) and affect dimensions (averaged daily-level positive and negative affect). Daily sleepiness (KSS) did not adversely impact NHS nurses’ attentional or processing speeds (CRT). Fast rotating night shift exposure was associated with greater frequency of cognitive errors.

### Clinical Characteristics

At study entry, over two thirds of NHS nurses reported feeling sad or anxious the prior month (GSAQ) and over a third had symptoms of subthreshold or mild depression (PHQ-9). Around three quarters of nurses reported moderate to high stress levels (PSS- 10) and one in four met burnout criteria (MBI-HSS MP). Sleep difficulties were also prevalent: over half of nurses reported severe circadian rhythm disturbances (BRIAN); over three quarters reported clinically relevant sleep impairments (PSQI) and insufficient sleep due to work (GSAQ; an indirect indicator of potential shift work sleep disorder ‘SWSD’); insomnia-related symptoms were highly prevalent and nearly all nurses experienced one potential sleep disorder symptom (*at least sometimes*) over the preceding month (Insomnia, obstructive sleep apnoea, restless leg syndrome, or parasomnia; GSAQ). These findings are consistent with prior evidence which show health and social care professionals experience significantly higher rates of work-related poor health (stress, burnout, depression and anxiety) compared with other occupational groups (16). Poor mental health, depression, and work-related stress, for example, are highly prevalent among nursing staff, with these risks further amplified by sleep deprivation and night shift work patterns (11, 17).

Daily sleep monitored over two-weeks revealed NHS nurses working fast rotating shifts (on average) subjectively slept for less (8 minutes; TST), took longer to fall asleep (10 minutes; SOL), and had worse sleep efficiency (SE) compared with day shift nurses (unadjusted descriptive scores in **Supplemental File, Table S7**). Poorer sleep among NHS nurses on fast rotating shifts was further reflected by greater sleep disturbance scores reported over the preceding month (PSQI; **Supplemental File, Table S8**).

Differences in mean daily sleep diary indices (TST, SOL, SE), however, were minimal between day shift nurses and nurses working fast rotating shifts over the two-week study period. These sleep patterns are consistent with previous nursing studies and may partially reflect compensatory sleep from NHS nurses on fast rotating shifts via prolonged daytime sleep or extra sleep opportunities during off-duty days (10).

## Daily Smartphone-based ESM

### Daytime Mood or Affect on Sleep

Daytime mood and affect (positive or negative) was not associated with subjective nightly sleep outcomes (TST, SQ, SE). Whilst previous shift work ESM studies found poorer mood was linked to shorter next-day sleep (TST) and higher positive affect was associated with better sleep quality (SQ), the broader literature suggests that sleep is a stronger predictor of subsequent daytime affective experiences than vice versa (12). In the *EClocker Study*, NHS nurses reporting better daily mood fell asleep faster (shorter SOL times), an association not previously reported in ESM studies with shift workers (12). Emotion regulation (DERS-SF) was modelled as a confound but emerged as an independent trait that was positively associated with sleep parameters (TST, SQ, SOL, SE) albeit with very small coefficients. We also explored the impact of emotion regulation (DERS-SF) on daily positive and negative affect dimensions and found small associations. NHS nurses therefore with higher emotional dysregulation (DERS-SF) were more likely to have variability in positive and negative affect via smartphone-based ESM.

### Sleep on Next-Day Mood or Affect

Subjectively reported sleep did not impact next-day mood or affect (positive and negative), aside from perceived sleep quality. NHS nurses therefore with higher perceived sleep quality had better next-day mood, a finding consistent with previous research (12) whereby subjectively perceived sleep (i.e. individual sleep introspection) appears to modulate mood states (18). Better sleep quality was also (inversely) associated with a small magnitude increase in next-day negative affect; these findings were particularly unexpected given the sleep quality-mood associations (**Table 3**) but may reflect domain measurement. Mood, for example, was averaged from a polarised scale (1 ‘lowest’ to 10 ‘highest’) whereas affect dimensions were composite scores of four momentary positive (PA) or six negative (NA) unipolar Likert scale items.

### Sleep on Next-Day Cognition

Nurses’ momentary sleepiness (KSS) did not impact attention or reaction time processing speeds (CRT). These findings may be attributable to study design constraints, since sleepiness was not rated concomitantly with the cognitive attentional task. A recent meta-analysis in healthy adults found one night of reduced sleep (after sleep restriction) increased sleepiness and impaired sustained attention – however, there were no sleep effects (as with our study) on Choice Reaction Time (19). Shorter, more ‘complex’ cognitive tasks such as the CRT may be less sensitive to sleep loss or fatigue due to compensatory prefrontal processes – these neural circuits may aid completion of more difficult, arousing tasks or maintain performance despite sleep deprivation (19, 20).

Exposure to fast rotating shifts during the study appeared to worsen NHS nurses’ cognitive performance, with a positive association with the number of incorrect Choice Reaction Time (CRT) trial responses. There is a wide body of literature that documents impaired cognition and performance among sleep-deprived healthcare workers, particularly on psychomotor vigilance tasks (PVTs) which share similar reaction-based performance assays with the CRT, including reduced cognitive response speeds and more frequent attention lapses. It is difficult to draw firm conclusions, nonetheless, and further analyses are required to determine whether this is a long-term dose-response relationship or the product of one or multiple contiguous night shifts. Fatigue and circadian-induced cognitive impairment are compounded by long work hours, night shift rotations, and reduced recovery opportunities which could jeopardise patient safety, quality of care, and increase clinical risk for errors and accidents (e.g., occupational, medication, or patient care errors) (8, 20). The insidious effects of fatigue and cumulative sleep deprivation are often underestimated by healthcare staff, despite their adverse impacts, including compromised decision-making, increased susceptibility for errors and cognitive lapses, and diminished performance in safety-critical settings.

## Future Research

Mood outcomes were averaged daily across multiple ratings. Secondary analyses could categorise mood responses based on time of day. Prior research, for example, has found emotional states are evaluated differently depending on the sampling prompt time (21). A recent longitudinal study with medical interns (22) also found daily mood scores naturally cycled based on the circadian clock phase, with lowest mood reported early in the morning (close to the circadian nadir at around 5 A.M.) and mood deteriorating with longer time spent awake. Diurnal mood variation is also a core feature of clinical depression - and for adults with high depressive symptomology - with worse morning mood and better rated evening mood (6, 23). These diurnal mood extremes can be induced by circadian dysregulation, and patterns are typically reversed in healthy individuals (i.e. worsening of evening-mood and declining positive affect). Our study sampled mood at multiple intervals to mitigate potential time-of-day influences – however, since mood was averaged in GAM models, further analyses could control for mood timings in relation to sleep characteristics. There is also limited data on specific risks of sleep-driven mood perturbations across shifts and over time (24, 25).

Time-lagged models could explore temporal dynamics between day-to-day sleep and mood variables over the wider two-week ESM period. Rather than assessing proximal next-day sequential associations, future analyses could examine shift workers sleep on one given night, predicting future sleep changes or affective symptoms over subsequent days or later time windows. In clinical populations, for example, better sleep quality predicted greater positive affect in psychosis over a 5-day lagged window (26).

Greater intraindividual sleep variability, lower rhythmicity, and more irregular sleep-wake patterns are associated with poorer mood and depressive symptoms (27, 28). Future work could examine NHS nurses’ day-to-day sleep-wake timing consistency, sleep regularity, sleep opportunity and recovery (including quick returns), schedule consistency and shift characteristics and its impact in driving detrimental mood symptomatology over the two-week period.

While this work predominantly focused on sleep diary ratings (notably sleep quality and sleep satisfaction) alongside daily ESM and questionnaire-based markers of subjective sleep experiences; a second phase of analyses will investigate the alignment with actigraphy-derived sleep measures (ActiGraph wGT3X-BT), sleep regularity, intraindividual sleep variability and dynamic time-lagged associations. This is especially important given the well-documented discrepancies between self-reported sleep patterns and objectively measured sleep metrics (12, 18). Findings from the *EClocker Study* will also provide an important foundation for future sleep-circadian research, including the *CHiP-D Study* [227099/Z/23/Z] which explores sleep-circadian disturbances, mental health, and cognitive outcomes among healthy volunteers, NHS shift workers, and patients with psychosis or depression.

A similar proportion of NHS nurses completed daily sleep-mood smartphone-based ESM monitoring across astronomical spring and summer (51%), compared with autumn and winter (49%). Future work, however, could control for seasonal shifts as a covariate in the GAM models or conduct repeated ESM sampling across seasons, as previously recommended (29). Additionally, nurses’ work shift patterns, sleep timing, and mood could be examined in relation to aberrant light-dark exposure including the pattern of light timing, intensity, duration and spectrum. A comparative sample of night shift-working nurses monitored across three seasons in the UK and Germany (30) found highly variable light exposure profiles, even for nurses working the same shifts within the same hospital. These light exposure differences (relative to individual circadian phase) appear to strongly influence circadian adaptation to night shift schedules in healthcare staff.

## CLINICAL IMPLICATIONS

The *EClocker Study* is the first to employ real-time digital sampling monitoring to evaluate dynamic, reciprocal associations of nightly sleep and daily affective experiences in NHS healthcare staff. Importantly, multidimensional affective constructs were assessed daily - including mood, emotions, and affect – along with clinically-relevant sleep indices, revealing complex sleep-affective dyad patterns. Preliminary evidence suggests that prioritising sleep quality could also serve as a therapeutic target to optimise mood symptoms in frontline NHS workers.

## Data Availability

All data produced in the present study are available upon reasonable request to the authors

## SUPPLEMENTARY FILES

Supplemental File 1

## FOOTNOTES

### Contributors

Conceptualisation (RH, TCD, SS); Methodology (RH, TCD, SS) with advice on cognitive task methodology from (AH, PJH); Data collection (RH); Formal analysis (RH, DWJ, NG, ZC); Writing - original draft preparation (RH); Writing - review and editing (all authors); Supervision (TCD, SS). All authors reviewed and approved the final version of the manuscript.

## Funding

This paper represents independent research [part] funded by the National Institute for Health and Care Research (NIHR) Maudsley Biomedical Research Centre at South London and Maudsley NHS Foundation Trust and King’s College London. The views expressed are those of the author(s) and not necessarily those of the NIHR or the Department of Health and Social Care.

This work is partly supported by the Wellcome Trust [CHiP-D Study: Grant Number 227099/Z/23/Z] for authors RH, DJ, AH, SS, TD

## Competing interests

AH is cofounder and codirector of H2 Cognitive Designs, a company that licenses online assessment technology for research and healthcare purposes. AH is also founder and director of Future Cognition Ltd, a company that develops cognitive assessment technology for third parties. PJH is also cofounder and codirector of H2 Cognitive Designs. The remaining authors declare no competing interests.

## Provenance and peer review

Not commissioned; externally peer reviewed.

## Author declaration

This article incorporates preliminary material revised and reworked from Doctoral thesis work [submitted by author Robert Hickman for the Doctor of Philosophy at King’s College London].

## Data availability statement

Data are available upon reasonable request.

## Ethics statements

Patient consent for publication

Not applicable.

Ethical approval: This study involves human participants and was approved by the Research Ethics Committee at King’s College London (HR-19/20-17792). The study was also approved by the Health Research Authority (HRA) and all participating NHS Trusts, Integrated Research Application System (IRAS) (289592). NHS organisations operated as Participant Identification Centres (PICs). Participants gave informed consent to participate in the study before taking part.

## Acknowledgements

The authors thank the research participants, NHS staff, and NHS Foundation Trusts who contributed their time and efforts towards this research.

## Notes

### Author Declarations

Ethical approval: This study involves human participants and was approved by the Research Ethics Committee at Kings College London (HR-19/20-17792). The study was also approved by the Health Research Authority (HRA) and all participating NHS Trusts, Integrated Research Application System (IRAS) (289592). NHS organisations operated as Participant Identification Centres (PICs). Participants gave informed consent to participate in the study before taking part.

