## Supplemental File 1 for "Temporal Dynamics of Daily Sleep, Mood, and Cognition in NHS Shift Workers: A Digital Experience Sampling (ESM) Study"

### Temporal Dynamics of Daily Sleep, Mood, and Cognition in NHS Shift Workers: A Digital Experience Sampling (ESM) Study

Robert Hickman, Dan W. Joyce, Nicholas Gray, Adam Hampshire, Peter J. Hellyer, Ziyuan Cai, Sukhi Shergill, Teresa C. D'Oliveira

#### TABLE OF CONTENTS

PARTICIPANT FLOW DIAGRAM
SCREENING MEASURES
DAILY MONITORING AND SMARTPHONE-BASED ESM MEASURES
NURSING SHIFT PATTERNS
RETROSPECTIVE MEASURES
SUPPLEMENTAL RESULTS, SAMPLE SIZE DETERMINATION, DATA CLEANING

#### PARTICIPANT FLOW DIAGRAM

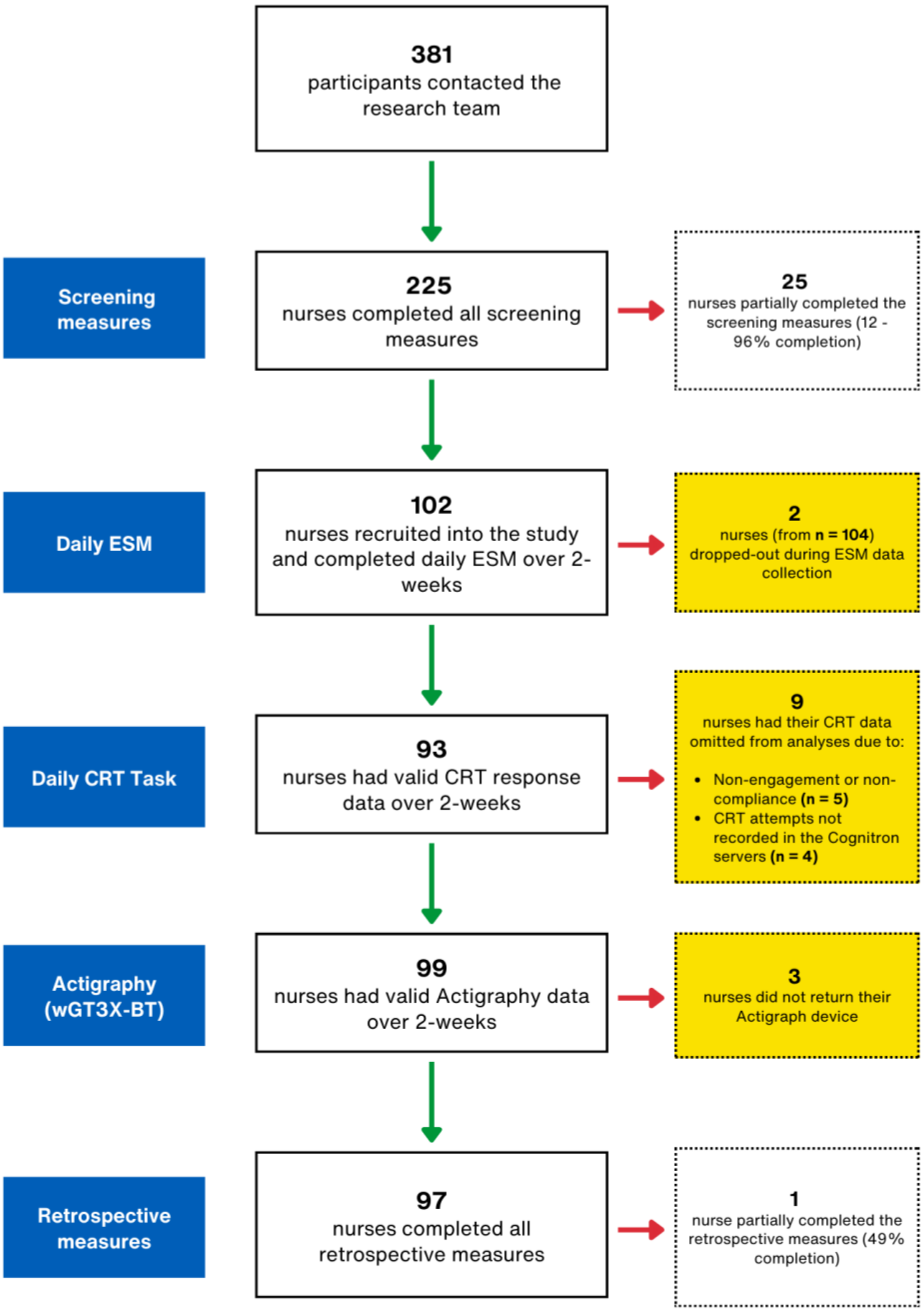


**Figure S1.** Participant Flow Diagram and data completion at each stage of the *EClocker Study* in Phase 1

Recruitment involved print and digital media outreach, mailing lists, and social media. Study information was shared with senior nursing leadership at participating NHS

Foundation Trusts and cascaded down to NHS staff. All participants provided informed consent online before taking part in the study.

#### SCREENING MEASURES

*EClocker Study* measure descriptions are provided below, with amended details from the original IRAS Study Protocol (V3.2; IRAS 289592).

##### Patient Health Questionnaire (PHQ-9)

The PHQ-9 is a 9-item diagnostic tool to screen for depression with scores ranging from 0-27 [1-4]. A systematic review concluded the PHQ-9 had, "acceptable diagnostic properties for detecting major depressive disorder for cut-off scores between 8 and 11" [2]. According to NICE guidelines, PHQ-9 scores less than 16 encompass both subthreshold and mild depression [5].

##### Mood Disorder Questionnaire (MDQ)

The MDQ [6] is a screening instrument for bipolar disorder. Three sections assess bipolar I disorder (BDI), bipolar II disorder (II), and bipolar disorder-not otherwise specified (BD-NOS) according to the Diagnostic and Statistical Manual of Mental Disorders Fourth Edition [7 8]. An MDQ optimal cut-off score of 6 [9 10] or ≥7 [11 12] has previously been utilised. Dimensions of bipolar disorder are considered for MDQ scores ≥7 (Items 1-13) with further clinical assessment needed if combined with symptom clustering (Item 14) and functional impact problems (Item 15).

##### Global Sleep Assessment Questionnaire (GSAQ)

The GSAQ [13] is a brief, 11-item screener of insomnia, obstructive sleep apnoea, restless leg syndrome / periodic limb movement, parasomnias, excessive daytime sleepiness (EDS) and impaired daytime function. Mood, life activities, and sleep symptoms over the previous four weeks are reported on a three-point scale with total scores ranging from 15-60. A systematic review identified the GSAQ as the, "most suitable instrument" for a general screener of multiple sleep disorders [14] and has previously been used to assess healthcare workers [15]. Symptoms were based on GSAQ Items 1-9 (excluding work-related Item 4) and responses were categorised as '*at least sometimes'* from the cumulative responses of '*Sometimes, Usually,* or *Always*' [in line with prior studies e.g., 16].

##### The Caen Chronotype Questionnaire (CCQ)

The (original) CCQ [17-19] is a 16-item measure of two chronotype dimensions; Morningness-Eveningness (CCQ-ME; 8-items) and Amplitude (CCQ-A; 8-items). A reduced two-factor model was also proposed by Hickman, et al. [20] as the most parsimonious and had best overall fit (4 ME items; 5 DI items).

#### The Morningness-Eveningness Questionnaire (MEQ)

The MEQ [21] is a measure of chronotype and has previously been validated with actigraphic data [22]. The MEQ consists of 19 items to assess individual differences in Morningness and Eveningness (ME). MEQ scores range from 16-86; ≤41 = 'evening types', ≥59 = 'morning types'; 42-58 = 'intermediate types'.

#### Biological Rhythms Interview of Assessment in Neuropsychiatry (BRIAN)

The BRIAN [23] is an 18-item tool to assess biological rhythm domains with scores ranging from 18-72; sleep activities, social rhythms, and eating patterns. Chronotype domain items 19-21 are not included in the BRIAN Total score. The BRIAN has been validated with objective markers of circadian rhythmicity including sleep-activity patterns from actigraphy and urinary melatonin metabolites [24-26]. Prior studies [27 28] identified Delayed Sleep-Wake Phase Disorder (DSWPD) from BRIAN total scores (≥40) in a Japanese sample (J-BRIAN-SR). BRIAN Total scores (>40) have also been used as a threshold for circadian rhythm alterations in bipolar which predicted depressive symptoms, suicidal risk, and emotion dysregulation [29].

#### Perceived Stress Scale (PSS-10)

The PSS-10 [30] is an instrument used to appraise perceived stress. A global stress score is generated from 10 items which ask about thoughts and feelings during the last month with scores ranging from 0-40. Although the PSS is not a diagnostic instrument and has no validated cut-offs, previous studies with nursing staff [e.g., 31 32 33] and international samples [34] have categorised PSS scores for moderate (14-26) and high (27-40) stress levels.

### DAILY MONITORING AND SMARTPHONE-BASED ESM MEASURES

Nurses downloaded the ExpiWell app onto their smartphone directly. The ExpiWell platform is GDPR-compliant and compatible with both iOS and Android devices. ExpiWell (formerly 'ExpiMetrics') has been widely used, with prior research (as with the *EClocker Study*) collecting daily sleep outcomes [35 36], mood, affect, or cognition with the smartphone platform [37-40].

#### Consensus Sleep Diary (CSD)

An adapted version of the Consensus Sleep Diary [CSD; 41] collected day-to-day self-reported sleep outcomes. The CSD is a standardised, prospective diary incorporating recommended clinical sleep indices such as TST, SE, SOL, WASO, TIB and SQ or satisfaction [42]. At the end of the sleep diary, mood valence was rated on a scale from 1 (Lowest) to 10 (Highest), adapted from Fang, et al. [43] and developed by Foreman [44]. Nurses were prompted (as per the general CSD instructions) to complete the sleep diary preferably within one hour of getting out of bed. In line with prior research [e.g., 45], sleep diaries were scheduled early in the morning (05:30 AM; see **Table S2**) and were available for the remainder of that day to maximise response and completion rates.

#### Affective Events Diary (AED)

An adapted Affective Events Diary (AED) was completed daily [46]. Relevant emotional work events are identified from 12 discrete categories (e.g., conflict between others), with the opportunity to provide a brief description of each event (open-ended format). To aid completion, examples of relevant emotional events were provided and adapted from Junça-Silva, et al. [47], Mignonac and Herrbach [48], and Ohly and Schmitt [49]. Self-report items related to daily napping, alcohol use, caffeine consumption, physical activity and sleep medication were also reported [adapted from the expanded Consensus Sleep Diary for Evening, CSD-E; 41] along with the type of day (e.g., work, day off, or vacation). At the end of the AED, mood valence (as for the sleep diary) was rated on a scale from 1 (Lowest) to 10 (Highest), adapted from Fang, et al. [43] and developed by Foreman [44].

#### NASA Task Load Index (NASA-TLX)

The NASA-TLX [50] is a reliable instrument of overall subjective workload. The NASA-TLX has previously been utilised in healthcare settings [51-53] and consists of six workload subscales: Mental Demand, Physical Demand, Temporal Demand, Performance, Frustration and Effort. Nurses in the *EClocker Study* reported their perceived workload towards the end of each work shift. The daily workload ESM item (NASA-TLX) was not included in final analyses and outcomes for this measure are not reported in this manuscript.

#### Karolinska Sleepiness Scale (KSS)

The 9-point KSS [54] evaluated subjective and situational sleepiness. It has previously been validated with electroencephalographic (EEG) and behavioural activity [55 56].

#### Affect

Momentary positive and negative affect items (e.g., 'Right now, I feel down') which have previously been used in the RADAR-MDD study [57-59] and PsyMate ESM clinical trials [60] were adapted (see **Table S1**). Affect items were comparable to previous sleep-affect and ambulatory ESM studies [e.g., 45 61 62-64] reported from Hickman, et al. [65] and based on standardised affective state measures such as the Positive and Negative Affect Schedule (PANAS-X, PANAS-SF), Profile of Mood States (POMS; POMS-SF) and Circumplex Model of Affect. The presentation order of affect items was fully randomised at each ESM push-notification.

#### Attentional Function (CRT)

The Choice Reaction Time (CRT) broadly assessed daily attentional function, reaction time performance, and processing speed. The CRT is a short 2-minute cognitive task developed by Cognitron (H2 Cognitive Design Ltd) and was delivered via the ExpiWell smartphone app using a secure link. The CRT task description has previously been described elsewhere and has a high test-retest reliability [66].

> *A black arrow either pointing left or right appeared on-screen, indicating the side of the screen the participant needed to click as fast as possible. The participants were presented with 60 stimuli with a 50% chance of each stimulus pointing left. The ISI [inter-stimulus interval] was jittered using a uniform random distribution between 0.5 and 2s.* (p. 8) [p.8 66]

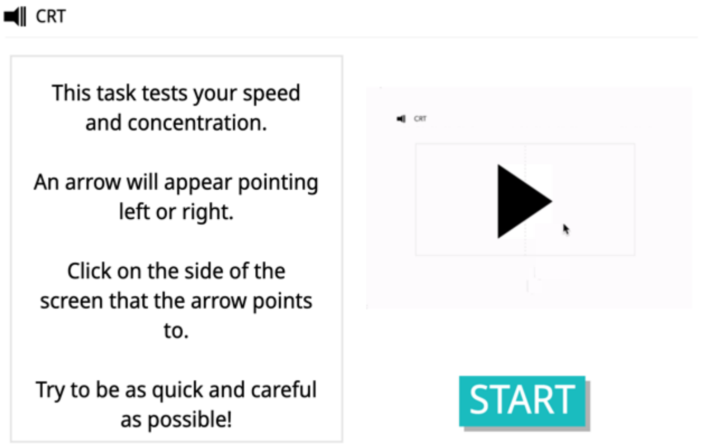


**Figure S2.** Choice Reaction Time (CRT) task developed by Cognitron (H2 Cognitive Design Ltd) was completed daily via the ExpiWell smartphone app

**Table S1** Daily Diary Questionnaires and Experience Sampling (ESM) Measures for the *EClocker Study*

| **Variable** | **Scale Option** |
|---|---|
| **Consensus Sleep Diary (adapted CSD)** | |
| **Sleep Diary**<br>A sleep diary is designed to gather information about your daily sleep pattern. Please complete your sleep diary every day. If possible, the sleep diary should be completed within **one hour** of getting out of bed. You should not worry about giving exact times. Just give your best estimate.<br><br>**What do the words 'bed' and 'day' mean on the diary?**<br>This diary can be used for people who are awake or asleep at unusual times. In the sleep diary, the word 'day' is the time when you choose or are required to be awake. The term 'bed' means the place where you usually sleep. | Sleep diary instructions |
| 1. What time did you get into bed? *This may not be the time that you began "trying" to fall asleep.* | Dropdown (timestamp) |

| | |
|---|---|
| 2. What time did you try to go to sleep? *Record the time that you began 'trying' to fall asleep.* | Dropdown (timestamp) |
| 3. How long did it take you to fall asleep? | Hour(s)<br>Minutes |
| 4. How many times did you wake up, not counting your final awakening? | 0 to ≥ 20 |
| 5. In total, how long did these awakenings last?<br>*For example, if you woke up 3 times for 20 minutes, 35 minutes, and 15 minutes, add them all up (20 + 35 + 15 = 70 min or 1 hr and 10 min).* | Hour(s)<br>Minutes |
| 6. What time did you finally wake up this morning? | Dropdown (timestamp) |
| 7. What time did you get out of bed for the day?<br>*This is with no further attempt at sleeping. It may be different from your final awakening time (e.g. you may have woken up at 6:35 a.m. but did not get out of bed to start your day until 7:20 a.m.)* | Dropdown (timestamp) |
| 8. How would you rate your sleep quality? | Very Poor<br>Poor<br>Fair<br>Good<br>Very Good |
| 9. Comments (if applicable)<br>*If you have anything that you would like to say that is relevant to your sleep, feel free to write it here. If not, please leave this blank.* | Open-ended response |
| On a scale of 1 (Lowest) to 10 (Highest), how is your **mood**? | 1 (Lowest) 2 3 4 5 6 7 8 9 10 (Highest) |
| What day of the week is it? | Dropdown (day of the week) |
| **Affect** | |
| Right now, I feel ... Cheerful | 0 (Not at all) 1 2 3 4 5 6 7 (Very much) |
| Right now, I feel ... Relaxed | 0 (Not at all) 1 2 3 4 5 6 7 (Very much) |
| Right now, I feel ... Content | 0 (Not at all) 1 2 3 4 5 6 7 (Very much) |
| Right now, I feel ... Hopeful | 0 (Not at all) 1 2 3 4 5 6 7 (Very much) |
| Right now, I feel ... Down | 0 (Not at all) 1 2 3 4 5 6 7 (Very much) |
| Right now, I feel ... Anxious | 0 (Not at all) 1 2 3 4 5 6 7 (Very much) |
| Right now, I feel ... Irritated | 0 (Not at all) 1 2 3 4 5 6 7 (Very much) |
| Right now, I feel ... Stressed | 0 (Not at all) 1 2 3 4 5 6 7 (Very much) |
| Right now, I feel ... Insecure | 0 (Not at all) 1 2 3 4 5 6 7 (Very much) |
| Right now, I feel ... Lonely | 0 (Not at all) 1 2 3 4 5 6 7 (Very much) |
| **Karolinska Sleepiness Scale (KSS)** | |
| Choose the number that best represents how **sleepy or alert** you feel right now | 1 = extremely alert<br>2 = very alert<br>3 = alert<br>4 = rather alert<br>5 = neither alert nor sleepy<br>6 = some signs of sleepiness<br>7 = sleepy, but no effort to stay awake<br>8 = sleepy, but some effort to stay awake<br>9 = very sleepy, great effort to stay awake, fighting sleep |
| Where am I **right now**? | At home<br>At family or friends place<br>At work<br>Transport<br>Somewhere else indoors |

| | |
|---|---|
| | Somewhere else outdoors |
| **NASA Task Load Index (NASA-TLX)** | |

**Instructions**
The evaluation you're about to perform is a technique that has been developed by NASA to assess the relative importance of six factors in determining how much workload you experienced while performing a task that you recently completed. These six factors are defined on the following page. Read through them to make sure you understand what each factor means.

**Definitions**

**Mental Demand** (low / high)
How much mental and perceptual activity was required (for example, thinking, deciding, calculating, remembering, looking, searching, etc.)? Was the task easy or demanding, simple or complex, forgiving or exacting?

**Physical Demand** (low / high)
How much physical activity was required (for example, pushing, pulling, turning, controlling, activating, etc.)? Was the task easy or demanding, slow or brisk, slack or strenuous, restful or laborious?

**Temporal Demand** (low / high)
How much time pressure did you feel due to the rate or pace at which the tasks or task elements occurred? Was the pace slow and leisurely or rapid and frantic?

**Performance** (good / poor)
How successful do you think you were in accomplishing the goals of the task set by the experimenter (or yourself)? How satisfied were you with your performance in accomplishing these goals?

**Effort** (low / high)
How hard did you have to work (mentally and physically) to accomplish your level of performance?

**Frustration Level** (low / high)
How insecure, discouraged, irritated, stressed, and annoyed versus secure, gratified, content, relaxed, and complacent did you feel during the task?

**Rating Scales**
You'll now be presented with a series of rating scales. For each of the six scales, evaluate the task you recently performed. Each scale has two endpoint descriptors that describe the scale. Consider your responses carefully in distinguishing among the different task conditions and consider each scale individually.

| | |
|---|---|
| **Mental Demand**<br>How much mental and perceptual activity did you spend for this task? | Low (0) (1) (2) Medium (3) (4) (5) (6) High (7) |
| **Physical Demand**<br>How much physical activity did you spend for this task? | Low (0) (1) (2) Medium (3) (4) (5) (6) High (7) |
| **Temporal Demand**<br>How much time pressure did you feel in order to complete this task? | Low (0) (1) (2) Medium (3) (4) (5) (6) High (7) |
| **Performance**<br>How successful do you think you were in accomplishing the goals of the task? ***Note the location of the endpoints are different before answering.*** | Good (7) (6) (5) (4) Medium (3) (2) (1) Poor (0) |
| **Effort**<br>How hard did you have to work to accomplish your level of performance? | Low (0) (1) (2) Medium (3) (4) (5) (6) High (7) |

| | |
|---|---|
| **Frustration**<br>How insecure, discouraged, irritated, stressed, and annoyed were you during this task? | Low (0) (1) (2) Medium (3) (4) (5) (6) High (7) |
| **Affective Events Diary (adapted AED)** | |
| **Emotional Events Diary**<br><br>**1.** If an event **at work** or **during** your working day has had an emotional impact on you (positive or negative), please select any of the categories below which apply. You may select more than one category for a single event (e.g. if you had an argument with a work colleague, select 'Co-worker' **and** 'Conflict with Others').<br><br>**2.** For each work event that has impacted you, please provide a short description of **what happened** and **how you felt** on the next page.<br>*You can **click this PDF link** for descriptions of relevant work events for each category. These are only examples; you may include any event you consider relevant and describe it as you see appropriate.* | Co-worker<br>Technical problem<br>Management<br>High workload<br>Low workload<br>Patients<br>Conflict with others<br>Aversive work<br>Making mistakes<br>Personal problem<br>Goals<br>Physical problem<br>Other event (not listed)<br>No event impacted me |
| **2.** For each work event that impacted you, please provide a short description of **what happened** and **how you felt**.<br>If **<u>no</u> event** impacted you, please leave this blank. | Open-ended response |
| **Nap or Doze**<br>Have you **napped or dozed** today?<br>*A **nap** is a time you decided to sleep during the day, whether in bed or not in bed. **"Dozing"** is a time you may have nodded off for a few minutes, without meaning to, such as while watching TV.* | Yes / No |
| How many times did you **nap or doze** today and for how long? | Number of times napped or dozed<br>Hour(s)<br>Minute(s) |
| **Alcohol**<br>Have you had an **alcoholic** drink today? | Yes / No |
| How many **alcoholic** drinks did you have? | Number of alcoholic drinks |
| What time was your last **alcoholic** drink? | Dropdown (timestamp) |
| **Caffeine**<br>Have you had a **caffeinated** drink today? (e.g. coffee, tea, soft drink, energy drink) | Yes / No |
| How many **caffeinated** drinks did you have? | Number of caffeinated drinks |
| What time was your last **caffeinated** drink? | Dropdown (timestamp) |
| **Exercise**<br>Have you **exercised** today?<br>*Exercise includes 'moderate intensity' activity which raises your heart rate and makes you breathe faster and feel warmer (e.g. a brisk walk, hiking, dancing).* | Yes / No |
| How much **exercise** did you get today (in minutes)? | Number of minutes |
| What time of day did you **exercise**? Select all that apply. | Morning<br>Afternoon<br>Evening |
| **Sleep medication**<br>Did you take any over-the-counter or prescription medication(s) to help you **sleep tonight**? *This includes 'herbals'.* | Yes / No |

| | |
|---|---|
| For **each** different sleep medication you took **tonight** to help you sleep, please list the:<br>1. Medication name<br>2. Dose<br>3. Time taken<br>*Example: "Sleepwell 50 mg 11 pm"* | Open-ended response |
| **Mood**<br>On a scale of 1 (lowest) to 10 (highest), how is your **mood**? | 1 (Lowest) 2 3 4 5 6 7 8 9 10 (Highest) |
| **Type of Day**<br>Was today a **working** day? | Work<br>Off<br>Holiday or vacation |
| Where did you **work** today? | Your workplace (e.g. hospital or office)<br>Own home<br>Family, partner or friends' place<br>Other (please describe) |

##### Actigraphy

As part of the larger *EClocker Study*, the ActiGraph wGT3X-BT accelerometer (ActiGraph LLC, Pensacola, FL, USA) was worn by NHS workers on the non-dominant ankle, positioned due to infection control policies across participating NHS Trusts. Actigraphy device data is *not* reported in the current paper.

#### NURSING SHIFT PATTERNS

As part of the online screening questionnaire, NHS nurses reported their speciality, employment history (e.g., years of experience), and NHS work schedule (e.g., shift system, shift intensity, weekly work hours, direction of shift rotation, shift lengths, and exposure to shift system). Push-notifications and survey scheduling were tailored in the *EClocker Study* for each nurse according to their day-to-day shift pattern, which was recorded during the in-person device collection visit. NHS England advises that rosters are published 6-12 weeks in advance but NHS nurses often have (unforeseen) short notice rota changes e.g., to respond to service demands, balance staffing levels, or to fill short-term (unexpected) staff absence due to sickness (Royal College of Nursing) [67]. Nurses may also swap agreed shifts with other staff members or work overtime (e.g., bank shifts). Nurses' final schedules and work hours were therefore updated until the first ESM push-notification and amended (when possible) over the two-week study period.

An example of an adjusted daily ESM schedule is outlined in **Table S2** with survey windows and reminder prompts individualised to account for the nurse's work hours and day shift pattern – their complete 2-week rota is reported in **Table S3**. Shift information and work schedule dimensions (e.g., shift timing, duration, weekly work hours, direction of rotation) were recorded at baseline in accordance with international reporting consensus [e.g., 68 69 70].

The exact timings of survey responses were recorded by the ExpiWell platform and are presented in the last column of **Table S2**. Nurses could postpone a momentary ESM notification if they were unable to respond at that time. Each survey, however, had a specified time window to respond with fixed prompt reminders to aid completion. In line with prior research [45], sleep diaries were scheduled early in the morning (05:30 AM) and were available for the remainder of that day to maximise response and completion rates.

**Table S2** Tailored ESM schedule for a day-working NHS shift nurse with shift hours starting at 08:00 AM and ending at 18:00 PM. In this example, all ESM surveys scheduled for that day were completed by the participant

| ESM Variable | Initial Push-Notification Time | Survey Window Open | Survey Window Close | Survey Reminder Prompt | Survey Response Time |
|---|---|---|---|---|---|
| Sleep Diary + Mood item | 05:30 AM | 05:30 AM | 23:59 PM | 240 mins | 06:06 AM |
| Choice Reaction Time (CRT) task | Notification **randomly** sent between 10:00 AM and 16:00 PM | 14:19 PM | 16:19 PM | 45 mins | 14:19 PM |
| Affect (PA / NA) + Sleepiness item | Notification **randomly** sent between 10:00 AM and 16:00 PM | 14:31 PM | 16:31 PM | 45 mins | 15:16 PM |
| Affective Events Diary + Mood item | 21:30 PM | 21:30 PM | 23:59 PM | 60 mins | 21:51 PM |

*Note.* Reminder prompts were sent after the initial push-notification for incomplete surveys. Survey response times for this participant (final column) are exact, as recorded in the ExpiWell platform. The daily workload ESM item (NASA-TLX) was not included in final analyses and outcomes for this measure are not reported in this manuscript.

**Table S3** Two-week work schedule for a day shift NHS nurse in *EClocker Phase 1* working from 08:00 AM to 18:00 PM

| | | Shift Pattern | Work Start Time | Work End Time |
|---|---|---|---|---|
| **Week 1** | Day 1 | Day shift | 08:00 | 18:00 |
| | Day 2 | Day shift | 08:00 | 18:00 |
| | Day 3 | Day shift | 08:00 | 18:00 |
| | Day 4 | Off work / rest-day | — | — |
| | Day 5 | Day shift | 08:00 | 18:00 |
| | Day 6 | Off work / rest-day | — | — |
| | Day 7 | Off work / rest-day | — | — |
| **Week 2** | Day 8 | Day shift | 08:00 | 18:00 |
| | Day 9 | Day shift | 08:00 | 18:00 |
| | Day 10 | Day shift | 08:00 | 18:00 |
| | Day 11 | Day shift | 08:00 | 18:00 |
| | Day 12 | Off work / rest-day | — | — |
| | Day 13 | Off work / rest-day | — | — |
| | Day 14 | Off work / rest-day | — | — |
| | Day 15 | Day shift | 08:00 | 18:00 |

#### RETROSPECTIVE MEASURES

##### Pittsburgh Sleep Quality Index (PSQI)

The PSQI [71] is a 24-item measure of seven sleep components: sleep quality (SQ), sleep latency (SL), sleep duration (TST), habitual sleep efficiency (SE), sleep disturbances, use of sleep medication and daytime function. Nineteen PSQI items generate a Global PSQI score (0-21) with higher scores indicating poorer sleep quality over the past month. A cut-off Global PSQI score of >5 is indicative of poor sleep quality and clinically relevant sleep impairments [see Riemann, et al. [72], Buysse, et al. [73], Mollayeva, et al. [74], Morin, et al. [75] for PSQI threshold details and Kyle, et al. [76] for sleep efficiency scores].

##### Emotional Regulation Strategy Questionnaire at Work (ERSQ-W)

The ERSQ-W [46] is a 14-item questionnaire based on Gross' five dimensions of emotion regulation at work. ERSQ-W categories include two situation selection, two situation modification, five cognitive change, three attentional deployment and two response modulation strategies. Participants reported whether they used any of the 14 strategies at least once in the previous 2 weeks (adapted timeframe) and the frequency of each.

##### Difficulties in Emotion Regulation Scale Short Form (DERS-SF)

The DERS-SF [77 78] is an 18-item self-report measure of emotion regulation ability across six subscales: Non-acceptance of emotional responses, difficulty engaging in goal-directed behaviour, impulse control difficulties, lack of emotional awareness, limited access to emotion regulation strategies, and lack of emotional clarity. Subscales are summed to form an overall 'DERS-SF Total' (ranging from 18-90), with higher scores indicating greater difficulties in emotion regulation and no clinical cut-offs.

##### Emotion Reactivity Scale (ERS)

The ERS [79] is a 21-item, self-report measure of emotional reactivity across three domains: emotional sensitivity (8 items); emotional arousal / intensity (10 items); emotional persistence (3 items). Items are summed to form a total ERS score ranging from 0-84, with higher scores indicating greater levels of emotional reactivity.

##### Positive and Negative Affect Schedule (PANAS-GEN)

The PANAS-GEN [80] is a 20-item instrument of trait positive (10 items) and negative (10 items) affect terms. Participants rate how they feel in general 'on average' [81]. PANAS-GEN Total scores range from 10-50 for both PA and NA scales, with no standardised cut-off points.

##### Maslach Burnout Inventory – Human Services Survey for Medical Personnel (MBI-HSS MP)

The MBI-HSS MP [82] is a 22-item measure of burnout across three components: Emotional Exhaustion (EE), Depersonalization (DP), and low sense of Personal Accomplishment (PA). The MBI-HSS MP has previously been validated in frontline healthcare professionals including doctors, medical residents, and nurses [83]. Prior systematic reviews [83 84] highlight the lack of consensus and variability in defining burnout among healthcare professionals (doctors, nurses, and medical residents) from the MBI instrument. Scores derived from the MBI are also considered arbitrary by the developers (as of 2016) due to a lack of diagnostic validity. Nonetheless, the most prevalent MBI-HSS MP cut-off scores have been identified among healthcare workers [83 84]. Burnout in our study was defined as nurses with both high-risk Emotional Exhaustion (EE ≥ 27) and high-risk Depersonalization (DP ≥ 10) MBI-HSS MP scores.

##### Perceived Work Schedule Control and Recovery Activities

See **Table S4** for (non-validated) measures of Perceived Work Schedule Control and Recovery Activities.

**Table S4** Perceived Work Schedule Control and Recovery Activities (Retrospective Measure)

| Variable | Scale Option |
|---|---|
| **Work Schedule Control** | |
| How much input do you have into the number of hours you work each week? | None At All, A Little, A Moderate Amount, A Lot, A Great Deal |
| When an unexpected personal or family matter arises, I have the ability to modify my schedule | Strongly Disagree, Disagree, Neutral, Agree, Strongly Agree |
| I have the ability to change my schedule when I have family or personal business to take care of | Strongly Disagree, Disagree, Neutral, Agree, Strongly Agree |
| How often are people you work with willing to swap hours with you or cover for you when you need to take time off for a family or personal matter? | Never, Rarely, Every once in a while, Sometimes, Always |
| **Recovery Activities** | |
| Do you have any hobbies or perform any specific activities in your free time? | Yes / No |
| Please Describe: | |
| (a) The nature of the activity | Open-ended response |
| (b) How often per week do you perform these activities? | Open-ended response |
| (c) At what time of the day do you perform the activities (morning, afternoon, evening)? | Open-ended response |

| | | |
|---|---|---|
| (d) | Where do you perform these activities (e.g., gym, indoors, outdoors)? | Open-ended response |

#### Cognitive Battery

A larger cognitive battery (**Figure S3**) was administered at the end of both *EClocker Phase 1* and *EClocker Phase 2* following completion of daily data collection (ESM and Actigraphy). Data from this cognitive battery is *not* reported in the current paper. Tasks were developed by Cognitron (H2 Cognitive Design Ltd) and delivered online via Qualtrics using a secure link: Cognitive Flexibility Task (IDED); Learning Curve Task; Digit Span Task; and Stop Signal Task (SST). The cognitive battery was designed to assess NHS nurses' executive function, response inhibition (impulse control), working-memory, reasoning, attention and cognitive flexibility.

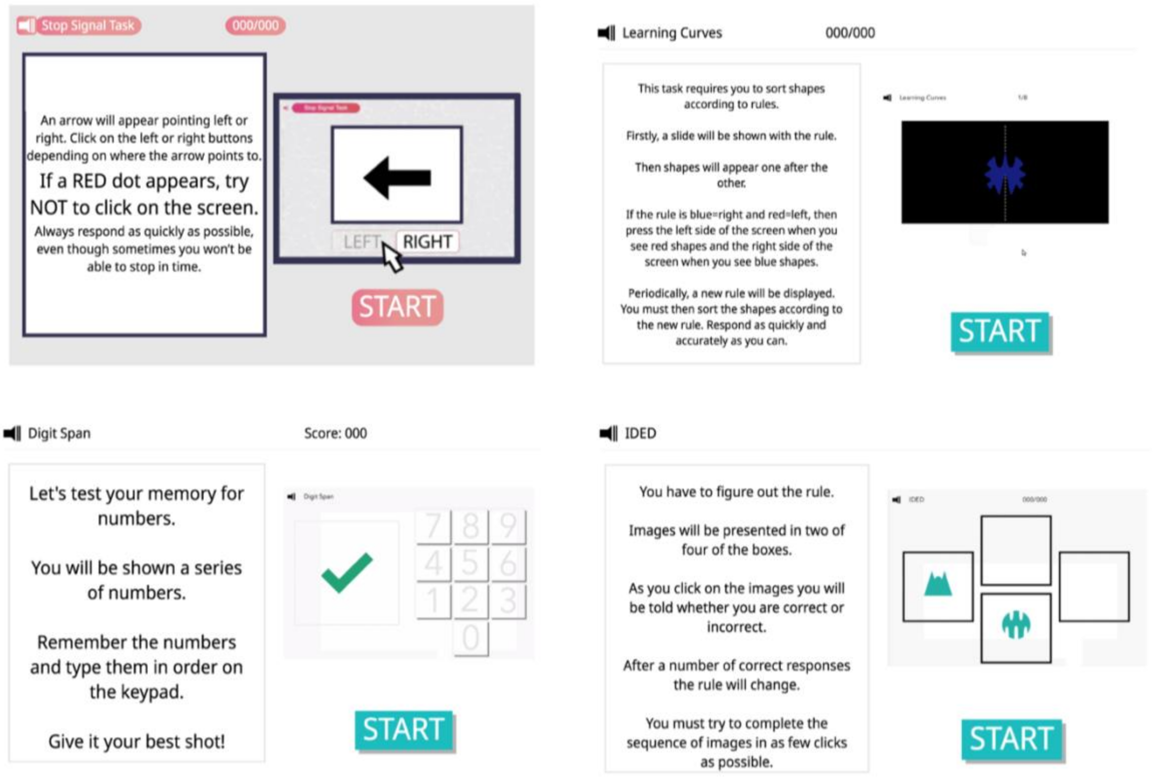


**Figure S3.** Stop Signal Task (SST), Learning Curve Task, Digit Span Task, Cognitive Flexibility Task (IDED) developed by Cognitron (H2 Cognitive Design Ltd) completed online via a secure Qualtrics link

#### SUPPLEMENTAL RESULTS

##### Sample Size Determination

An *a priori* calculation using G*Power software (version 3.1.9.6; http://www.gpower.hhu.de/) was conducted to determine the required sample size and to ensure the study was adequately powered. To test differences between two independent groups (NHS day shift nurses and nurses working fast rotating shifts), with a medium effect size (d=0.50), 80% power, and alpha rate of $\alpha = 0.05$, a sample size per group of n=50 was proposed.

##### Data Cleaning

*Sleep*

Sleep diary data cleaning was conducted using Python scripts to remove erroneous or implausible responses (see **Table S5** in **Supplemental File 1** for examples). Unresolvable participant sleep diary entry errors (for TST, SOL, SE) were conservatively removed from the dataset and subsequent GAM modelling.

*Gender*

The majority of NHS nurses were female (83.3%) which compares with staff demographics for NHS England (74% female) and the UK average for Registered Nurses (89% female) (NHS-England 2024 figures). Since the final sample was predominantly female, gender was removed as a covariate from the GAMs.

*Choice Reaction Time (CRT)*

Day-to-day CRT responses across two-weeks were pre-processed in Python for 93 nurses. Nine nurses (out of 102) had their CRT data omitted from final GAM analyses due to non-engagement or non-compliant behaviour with the study protocol (n = 5) or technical error with missing task data not retrieved from Cognitron servers (n = 4). Non-compliance included 4 participants who failed to interact with the CRT stimulus (trials with a 100% time-out rate - a sign of participant disengagement or no interaction with the stimulus) and 3 participants exhibiting a repetitive response pattern (consistently clicked the same screen location, 100% right or left across trials).

Definitions and calculations of each daily ESM self-reported variable are outlined in **Table S5**. Scoring and data cleaning of these daily ESM variables were in line with prior conservative approaches [85].

**Table S5** Definitions and calculations of each daily ESM self-reported variable (sleep, mood, and affect)

| Variable | Definition and/or Calculation of Indices | Data Cleaning |
|---|---|---|
| Total Sleep Time (TST) | Total time asleep was calculated from CSD sleep onset-offset timings, minus sleep latency (SOL) and wake after sleep onset (WASO) | TST (hours) not in the range of $2 \leq \text{TST} \leq 14$ were excluded. Implausible sleep-wake timing responses (e.g., impossible values due to participant error) were removed |
| Sleep Efficiency (SE) | $\mathrm{SE} = \frac{\mathrm{TST}}{\mathrm{TIB}}$ Percent of time asleep (TST) out of time spent in bed (TIB) | SE (%) not in the range of 0-100 were excluded. If TST was outside the range of 2-14 hours, subsequent SOL and SE data were also removed |
| Sleep Onset Latency (SOL) | The duration of time (minutes) participants reported it took them to fall asleep when they began 'trying' (after sleep attempt) | Implausible responses and common participant entry errors were excluded (e.g., SOL + WASO > sleep opportunity window). If TST was outside the range of 2-14 hours, subsequent SOL and SE data were also removed |
| Sleep Quality (SQ) | Perceived quality of sleep was rated from 1 (very poor) to 5 (very good). An open-ended section also allowed for free response entries: '*If you have anything that you would like to say that is relevant to your sleep, feel free to write it here'* | |
| Mood | Mood valence was rated on a scale from 1 (lowest) to 10 (highest). Mood scores were averaged across all mood survey responses that day | |
| Affect | 10 affect items (4 PA; 6 NA) were rated on a 7-point Likert scale (anchored at 0 = not at all, to 7 = very much)<br><br>Daily positive and negative affect items were each aggregated to form daily-level averages of positive or negative affect | |

**CSD** = Consensus Sleep Diary; **NA** = Negative Affect; **PA** = Positive Affect; **SE** = Sleep Efficiency; **SOL** = Sleep Onset Latency; **SQ** = Sleep Quality; **TIB** = Time Spent In Bed; **TST** = Total Sleep Time; **WASO** = Wake After Sleep Onset

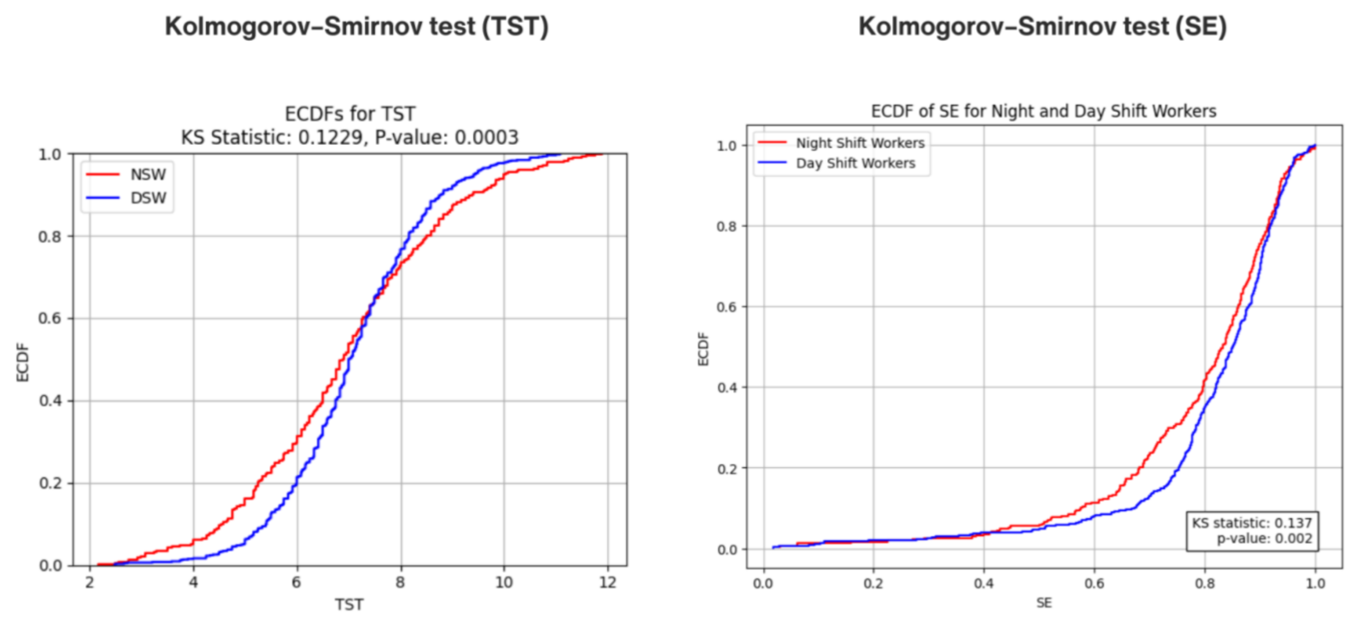


**Figure S4.** Empirical Distribution Functions (ECDF) and Kolmogorov–Smirnov Tests (Two-Sided) for total sleep time (left; TST) and sleep efficiency (right; SE) values for NHS nurses working fast rotating shifts (Red) and Day Shift Nurses (Blue). Sleep outcomes (TST and SE) are taken from the CSD sleep diaries during *EClocker Phase 1*

**Table S6** Spline Points for variables used in the General Additive Models (GAMs)

| **Variable** | **Spline Points** |
|---|---|
| Mood (CSD) | 4 |
| Mood (AED) | 6 |
| Mood (Averaged) | 6 |
| TST | 5 |
| SL | 5 |
| SQ | 5 |
| Affect (Averaged) | 5 |
| Affect (PA) | 5 |
| Affect (NA) | 5 |
| NSW | 4 |
| Age | 8 |
| PHQ-9 | 6 |
| MEQ | 4 |
| DERS-SF | 4 |

**CSD** = Consensus Sleep Diary; **AED** = Affective Events Diary; **TST** = Total Sleep Time; **SL** = Sleep Latency; **SQ** = Sleep Quality; **PA** = Positive Affect; **NA** = Negative Affect; **NSW** = NHS nurses working fast rotating shifts; **PHQ-9** = Patient Health Questionnaire; **MEQ** = Morningness-Eveningness Questionnaire; **DERS-SF** = Difficulties in Emotion Regulation Scale Short Form

**Table S7** Smartphone-based ESM variables self-reported by NHS nurses during *EClocker Phase 1*. Sleep, mood, and affect scores are averaged across two-weeks of data collection and split by modality of work

| Variables | All Nurses (N = 102) | | | | Day Shift Nurses (N = 49) | | | | Fast Rotating Nurses (N = 53) | | | |
|---|---|---|---|---|---|---|---|---|---|---|---|---|
| | Mean | SD | Min | Max | Mean | SD | Min | Max | Mean | SD | Min | Max |
| Total Sleep (TST) | 7.04 | 1.58 | 2.48 | 11.88 | 7.10 | 1.35 | 2.48 | 11.08 | 6.96 | 1.85 | 2.50 | 11.88 |
| Sleep Latency (SOL) | 21.58 | 27.88 | 0.00 | 325.0 | 17.46 | 22.10 | 0.00 | 240.0 | 27.17 | 33.42 | 0.00 | 325.0 |
| Sleep Efficiency (SE) | 0.82 | 0.13 | 0.28 | 1.00 | 0.83 | 0.12 | 0.28 | 1.00 | 0.80 | 0.13 | 0.38 | 1.00 |
| Sleep Quality (SQ) | 3.23 | 0.72 | 2.00 | 4.00 | 3.22 | 0.72 | 2.00 | 4.00 | 3.23 | 0.73 | 2.00 | 4.00 |
| Mood | 6.03 | 1.67 | 1.00 | 10.00 | 6.15 | 1.62 | 1.50 | 10.00 | 5.88 | 1.73 | 1.00 | 10.00 |
| Positive Affect (PA) | 1.93 | 1.01 | 0.00 | 5.00 | 1.92 | 0.97 | 0.00 | 5.00 | 1.94 | 1.05 | 0.00 | 5.00 |
| Negative Affect (NA) | 2.38 | 0.75 | 0.00 | 5.00 | 2.42 | 0.69 | 0.00 | 5.00 | 2.34 | 0.80 | 0.00 | 5.00 |

*Note.* Sleep parameters are from the daily CSD; **TST** in hours, **SOL** in minutes, **SQ** scored on a 1-5 scale; **Mood** scored on a 1-10 scale and averaged daily; **Affect** scored on a 0-7 scale and averaged daily for PA and NA. To compare day (DSW) and night shift (NSW) nurses, only sleep diaries completed the day after a nurse worked a 'standard day shift' or after days with no contracted work hours (non-working days) were included.

**Table S8** Summary scores for self-reported outcome variables (*EClocker Phase 1* Retrospective Measures). All NHS Nurses (N = 98), Day Shift Nurses (N = 49), Fast Rotating Shift Nurses (N = 49)

| Outcome Variable | All Nurses | | | | Day Shift Nurses | | | | Fast Rotating Nurses | | | |
|---|---|---|---|---|---|---|---|---|---|---|---|---|
| | Mean | SD | Min | Max | Mean | SD | Min | Max | Mean | SD | Min | Max |
| Sleep Quality (PSQI Global) | 7.2 | 3.3 | 2 | 16 | 6.5 | 2.9 | 2 | 15 | 8.0 | 3.5 | 3 | 16 |
| **Burnout (MBI-HSS MP)** | | | | | | | | | | | | |
| Emotional Exhaustion | 28.1 | 12.0 | 2 | 53 | 26.8 | 12.7 | 2 | 46 | 29.4 | 11.2 | 11 | 53 |
| Depersonalization | 7.7 | 5.5 | 0 | 27 | 6.6 | 5.2 | 0 | 25 | 8.7 | 5.7 | 0 | 27 |
| Personal Accomplishment | 34.6 | 7.0 | 15 | 47 | 34.9 | 7.7 | 15 | 47 | 34.2 | 6.3 | 20 | 45 |
| **Affective State (PANAS-GEN)** | | | | | | | | | | | | |
| Positive Affect (PA) | 33.8 | 7.2 | 16 | 50 | 34.0 | 7.4 | 20 | 50 | 33.7 | 7.1 | 16 | 47 |
| Negative Affect (NA) | 19.1 | 6.7 | 10 | 40 | 18.6 | 7.0 | 10 | 40 | 19.7 | 6.5 | 11 | 36 |
| Emotion Regulation (DERS-SF Total) | 39.5 | 11.5 | 20 | 75 | 38.6 | 11.5 | 20 | 70 | 40.3 | 11.6 | 20 | 75 |
| Emotion Reactivity (ERS Total) | 28.0 | 16.0 | 0 | 75 | 26.1 | 14.7 | 0 | 67 | 30.0 | 17.2 | 0 | 75 |

*Note*. Total number of responses may differ for each outcome variable e.g., due to missing or invalid responses. **PSQI =** Pittsburgh Sleep Quality Index; **MBI-HSS MP** = Maslach Burnout Inventory – Human Services Survey for Medical Personnel; **PANAS-GEN** = Positive and Negative Affect Schedule; **DERS-SF** = Difficulties in Emotion Regulation Scale Short Form; **ERS** = Emotion Reactivity Scale
